# Differential manifestation of type 2 diabetes in Black Africans and White Europeans with recently diagnosed type 2 diabetes: A systematic review

**DOI:** 10.1101/2024.03.26.24304917

**Authors:** Davis Kibirige, Ronald Olum, Andrew Peter Kyazze, Bethan Morgan, Felix Bongomin, William Lumu, Moffat J. Nyirenda

## Abstract

**Aims:** The clinical manifestation of type 2 diabetes (T2D) varies across populations. We compared the phenotypic characteristics of Black Africans and White Europeans with recently diagnosed T2D to understand the ethnic differences in the manifestation of T2D.

**Methods:** We searched Medline, EMBASE, CINAHL, Google Scholar, African Index Medicus, and Global Health for studies reporting information on phenotypic characteristics in Black Africans and White Europeans with recently diagnosed T2D.

**Results:** A total of 26 studies were included in this systematic review. Of these, 12 studies and 14 studies were conducted on 2,586 Black Africans in eight countries and 279,621 White Europeans in nine countries, respectively. Compared with White Europeans, Black Africans had a lower pooled mean age (49.4±4.4 years vs. 61.3±2.7 years), body mass index (26.1±2.6 kg/m^2^ vs. 31.4±1.1 kg/m^2^), and a higher pooled median glycated haemoglobin (9.0 [8.0-10.3]% vs. 7.1 [6.7-7.7]%). Ugandan and Tanzanian participants had lower markers of beta-cell function and insulin resistance when compared with four White European populations.

**Conclusion:** These findings provide evidence of the ethnic differences in the manifestation of T2D, underscoring the importance of understanding the underlying genetic and environmental factors influencing these phenotypic differences and formulating ethnic-specific approaches for managing and preventing T2D.

**HIGHLIGHTS:**

- Emerging evidence suggests differences in the presentation of type 2 diabetes in Black Africans and White Europeans.
- In this systematic review, we reported that compared with White Europeans, Black Africans presented with a lower mean age and body mass index, less co-existing hypertension, and more hyperglycaemia at the time of diagnosis of type 2 diabetes.
- Compared with some White European populations, Ugandan and Tanzanian participants presented with features of pancreatic beta-cell dysfunction and less insulin resistance.

## INTRODUCTION

Type 2 diabetes (T2D) is characterised by marked heterogeneity in the clinical presentation, pathophysiology, progression, and therapeutic response across populations [1]. This may be due to the differences in genetics and environmental exposures. For example, several studies have demonstrated striking differences in T2D phenotypes between Asian, mixed ancestry, and White populations of European ancestry [2-5].

In contrast, studies that have compared well-characterised adult Black Africans with other populations are lacking. Emerging data suggest that there may be differences in the manifestation of T2D in Black African and White populations of European ancestry [6]. This evidence is mainly derived from anecdotal observations and small clinical studies that lack detailed information on metabolic and immunologic characteristics of individuals [6-8]. However, understanding these phenotypic differences is important to guide the formulation of optimal targeted population-specific preventive and therapeutic approaches for T2D, especially in Black African populations.

To gain a better understanding of the differences in the manifestation of T2D in Black Africans and White Europeans, we systematically reviewed relevant original studies published between 1946 and February 2024. We compared specific demographic, clinical, anthropometric, and metabolic characteristics in these two distinct populations to establish if any phenotypic differences exist.

## MATERIALS AND METHODS

This systematic review was conducted according to the criteria outlined in the Preferred Reporting Items for Systematic Reviews and Meta-Analyses (PRISMA) statement [9]. The PRISMA checklist is available as a supplementary Table 1. The study protocol was registered in the PROSPERO International Prospective Register of Systematic Reviews (CRD42023416884).

### Search strategy

With the help of a librarian (BM), we searched Medline, EMBASE, CINAHL, Google Scholar, African Index Medicus, and Global Health for published studies from 1946 to 7^th^ February 2024. The following search terms were used: “recently diagnosed diabetes” OR “recent-onset diabetes” OR “recent-onset type 2 diabetes” OR “newly diagnosed diabetes” OR “newly diagnosed type 2 diabetes” OR “newly-diagnosed type 2 diabetes” OR “newly diagnosed patients with type 2 diabetes” OR “new-onset diabetes” OR “recently diagnosed type 2 diabetes” OR “recently diagnosed T2D” OR “recently diagnosed with type 2 diabetes mellitus” OR “recently diagnosed with type 2 diabetes” OR “newly diagnosed type 2 diabetes” OR “newly diagnosed T2D” OR “new-onset type 2 diabetes” OR “new-onset T2D” OR “adult-onset type 2 diabetes” AND Angola OR Benin OR Botswana OR ‘‘Burkina Faso’’ OR Burundi OR Cameroon OR ‘‘Cape Verde’’ OR ‘‘Central African Republic’’ OR Chad OR Comoros OR ‘‘Democratic Republic of Congo’’ OR Djibouti OR ‘‘Equatorial Guinea’’ OR Eritrea OR Ethiopia OR Gabon OR Gambia OR Ghana OR Guinea OR ‘‘Guinea Bissau’’ OR ‘‘Ivory Coast’’ OR ‘‘Cote d’Ivoire’’ OR Kenya OR Lesotho OR Liberia OR Libya OR Libya OR Madagascar OR Malawi OR Mali OR Mauritania OR Mauritius OR Morocco OR Mozambique OR Namibia OR Niger OR Nigeria OR Rwanda OR ‘‘Sao Tome’’ OR Senegal OR Seychelles OR ‘‘Sierra Leone’’ OR Somalia OR ‘‘South Africa’’ OR “South Sudan” OR Sudan OR Swaziland OR Tanzania OR Togo OR Uganda OR Zaire OR Zambia OR Zimbabwe OR ‘‘Central Africa’’ OR ‘‘West Africa’’ OR ‘‘Western Africa’’ OR ‘‘East Africa’’ OR ‘‘Eastern Africa’’ OR ‘‘Southern Africa’’ OR ‘‘sub Saharan Africa’’ OR ‘‘sub-Saharan Africa’’ OR Africa AND Albania OR Andorra OR Armenia OR Austria OR Azerbaijan OR Belarus OR Belgium OR “Bosnia and Herzegovina” OR Bulgaria OR Croatia OR Cyprus OR “Czech Republic” OR Denmark OR Estonia OR Finland OR France OR Georgia OR Germany OR Greece OR Hungary OR Iceland OR Ireland OR Italy OR Kazakhstan OR Kosovo OR Latvia OR Liechtenstein OR Lithuania OR Luxembourg OR Malta OR Moldova OR Montenegro OR Netherlands OR “North Macedonia” OR Norway OR Poland OR Portugal OR Romania OR Russia OR “San Marino” OR Serbia OR Slovakia OR Slovenia OR Spain OR Sweden OR Switzerland OR Turkey OR Ukraine OR “United Kingdom” OR England OR Scotland OR Wales OR “Vatican City”.

In addition to the search of the databases above, we also hand-checked the references of the articles whose full texts were retrieved for any additional studies.

### Study selection criteria

The preliminary screening of titles and abstracts to identify potentially eligible articles was done independently by two reviewers (DK and APK) after the duplicates were removed. Full texts of the potentially eligible studies were retrieved and reviewed for the information of interest. The inclusion criteria of studies were: cross-sectional studies, retrospective studies, cohort studies, case-control studies, and randomised controlled trials published between 1946 and 7^th^ February 2024 in the English language with any baseline information on the demographic (age at diagnosis, sex), clinical (self-reported history of hypertension, systolic, and diastolic blood pressure), anthropometric (body mass index [BMI] and waist circumference [WC]), and metabolic (fasting blood glucose [FBG], glycated haemoglobin [HbA1c], fasting C-peptide, lipid profile (total cholesterol, low-density lipoprotein cholesterol [LDLC], high-density lipoprotein cholesterol [HDLC], and triglycerides), homeostatic model assessment-2 insulin resistance [HOMA2-IR], and homeostatic model assessment-2 beta cell function [HOMA2-%B]) characteristics of Black Africans and White Europeans with recently diagnosed T2D. Because most of the phenotypic characteristics of interest (notably BMI, WC, insulin resistance, and pancreatic beta-cell function status) are altered by increasing duration of T2D, we restricted our selection to studies that enrolled participants within 18 months from the time of diagnosis of T2D.

In cases of any disagreements, an independent reviewer (FB) was consulted. We excluded case series and reports, studies published in languages other than English, and studies whose full texts could not be retrieved.

### Data extraction

After identifying the eligible studies, the relevant study information of interest was extracted using a Microsoft Excel 2016 form by one author (DK). The information that was extracted included: the last name of the first author and year of publication, country or countries where the study was conducted, number of study participants, number and proportion of female participants, proportion of participants with self-reported co-existing hypertension, mean ± standard deviation (SD) or median (interquartile range or IQR) of the age at diagnosis, systolic and diastolic blood pressures, BMI, WC, FBG, HbA1c, lipid profile (total cholesterol [TC], high-density lipoprotein cholesterol [HDLC], low-density lipoprotein cholesterol [LDLC], and triglycerides [TGL]), fasting C-peptide, HOMA2-%B, and HOMA2-IR.

### Operational definitions

We defined recently diagnosed T2D as a diagnosis of T2D made in the preceding 18 months. A Black African and White European participant was a participant residing in a particular African and European country, respectively, at the time the study was conducted. For the White European participants, we considered those classified as of White ancestry.

### Assessment of quality of studies

The quality of all eligible studies included in the systematic review was assessed using the modified Newcastle-Ottawa Scale (NOS) [10]. This was done independently by one author (APK).

### Study outcomes

The study outcome was the differences in the specific demographic, clinical, anthropometric, and metabolic characteristics between Black Africans and White Europeans with recently diagnosed T2D.

### Data analysis

Quantitative data analyses were conducted using STATA version 18.0 (StataCorp LLC, College Station, Texas, US). For continuous variables reported across two or more studies, we derived pooled estimates using two approaches: 1) a weighted mean of means was calculated to adjust for study sample sizes, and 2) a non-weighted median of medians was reported to provide a robust measure of central tendency less influenced by extreme values. To assess the pooled proportions for categorical variables (gender and co-existing hypertension), we employed meta-analytical techniques utilising the meta-command in STATA. Statistical heterogeneity among the included studies was quantified using the I² statistic. Additionally, potential publication bias was evaluated using Egger’s regression test and contour-enhanced funnel plots to facilitate the distinction between publication bias and other sources of asymmetry. A p<0.05 on Egger’s regression test indicated significant publication bias. All the quantitative analyses were stratified by race, namely, Black Africans and White Europeans. For single-study variables, where meta-analysis was not feasible, we provided a narrative synthesis.

### Ethical approval

Because this was a systematic review of published studies, no prior ethical approval was required.

## RESULTS

Figure 1 summarises the article selection in a PRISMA flow diagram.

The literature search returned a total of 776 articles. From these, 139 duplicates were removed. The titles and abstracts of the remaining 637 articles were reviewed and 72 articles were identified for full-text retrieval. Of the 72 articles, 54 were excluded and the remaining 18 were included in the systematic review. On hand-searching the references of the retrieved full texts, an additional eight articles were identified, making a total of 26 articles included in the systematic review.

**Figure 1.**
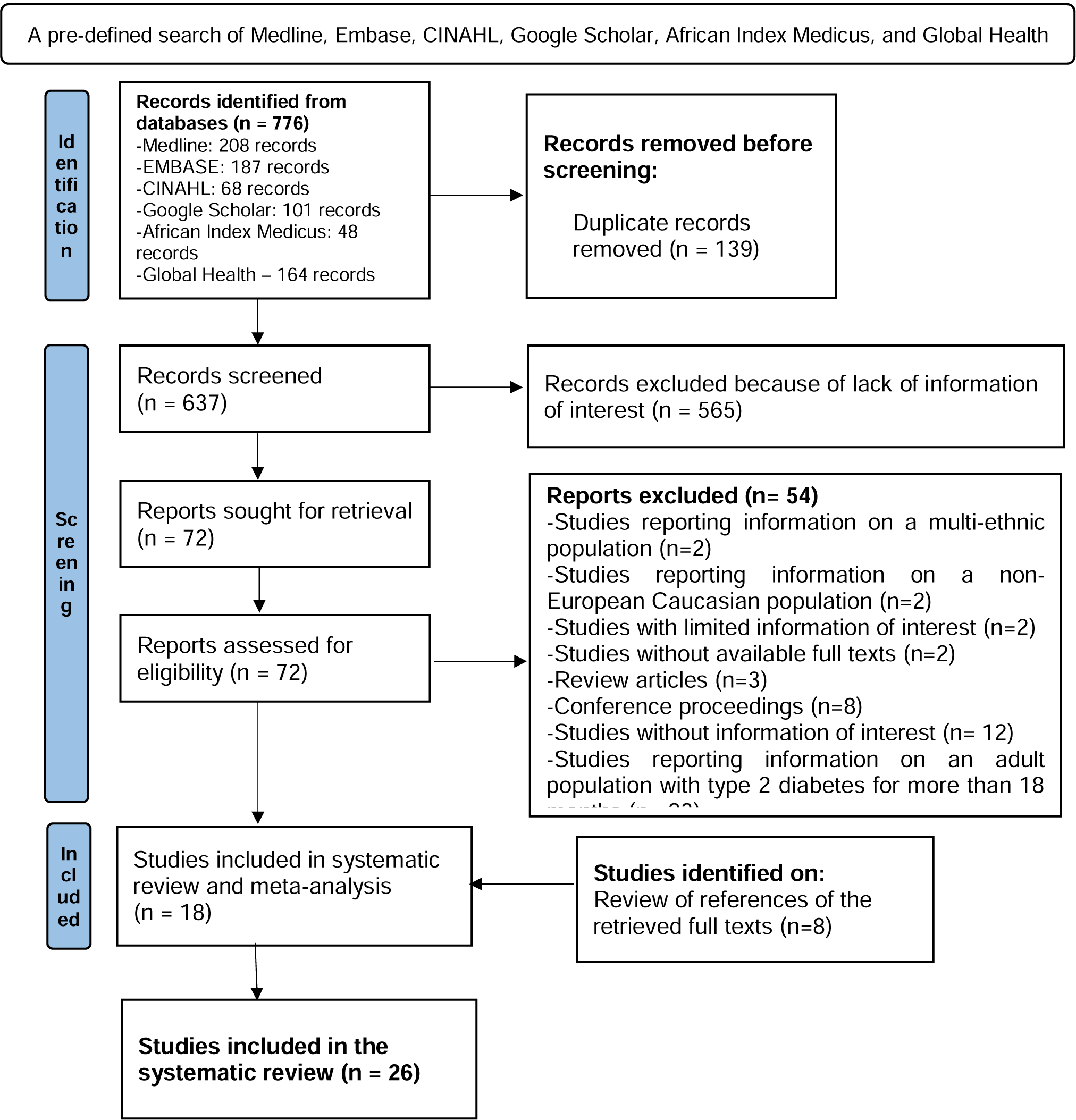
PRISMA flow diagram of selection of eligible studies.

### Characteristics of included studies

Tables 1 and 2 summarise the overall and specific characteristics of the studies included in the systematic review.

**Table 1.**
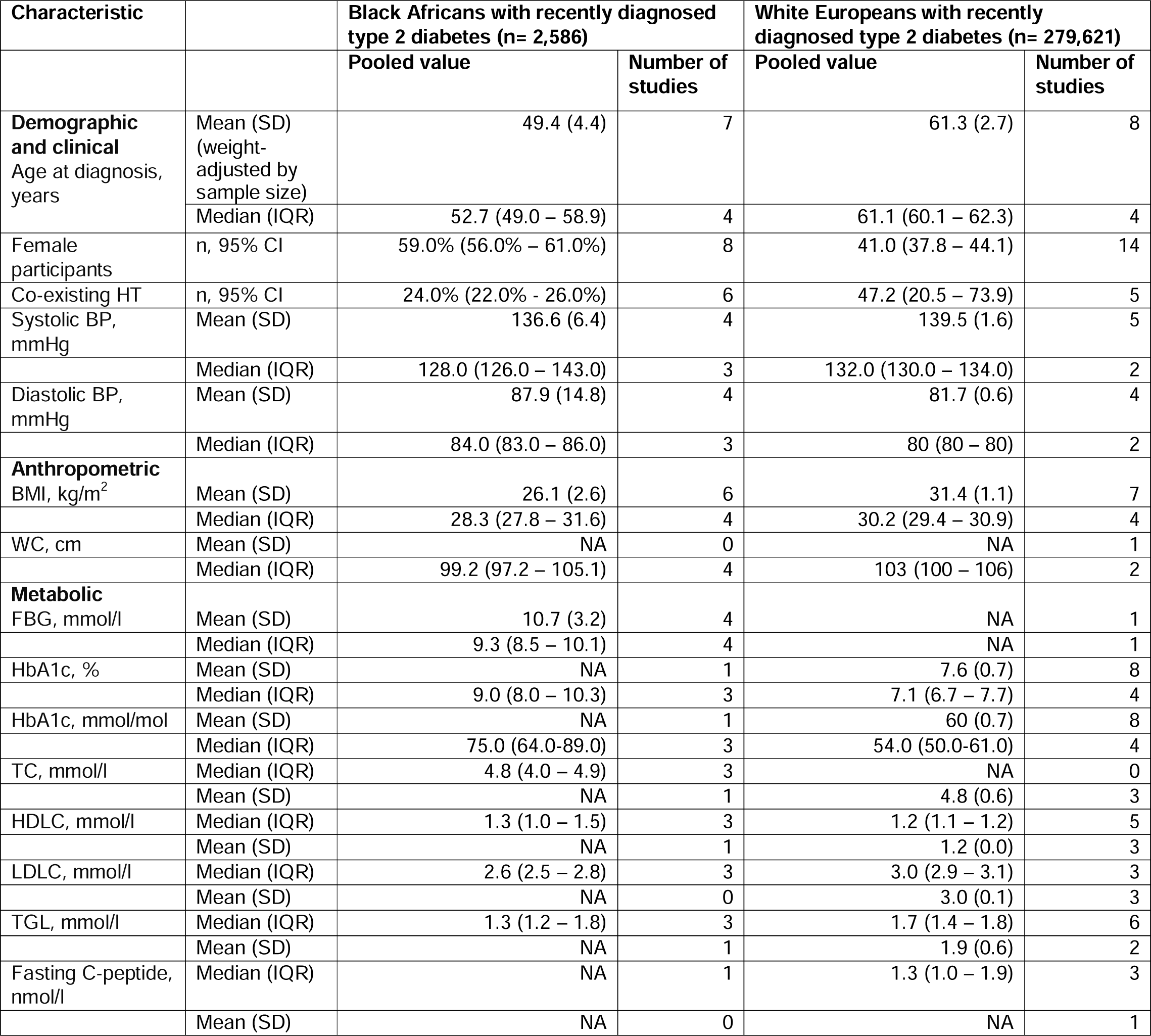
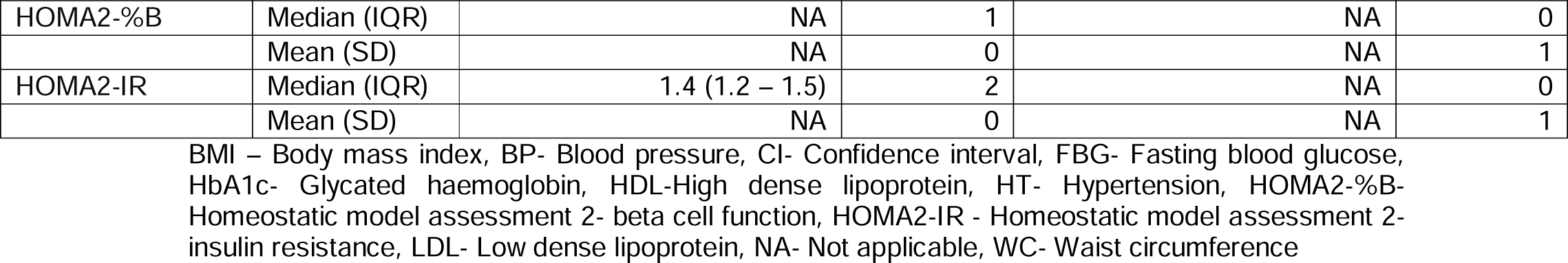
Overall characteristics of the studies included in the systemic review.

**Table 2.**
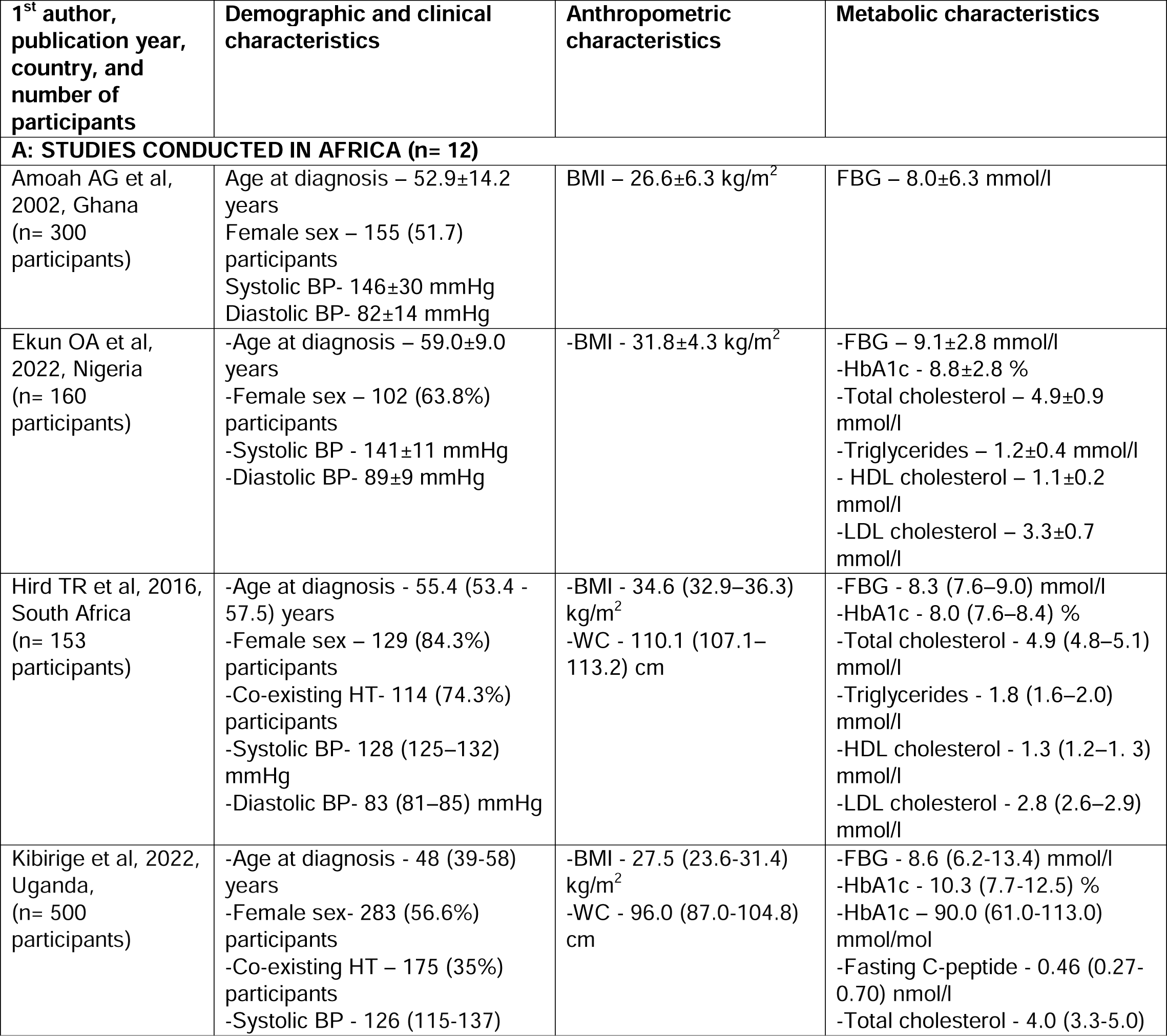
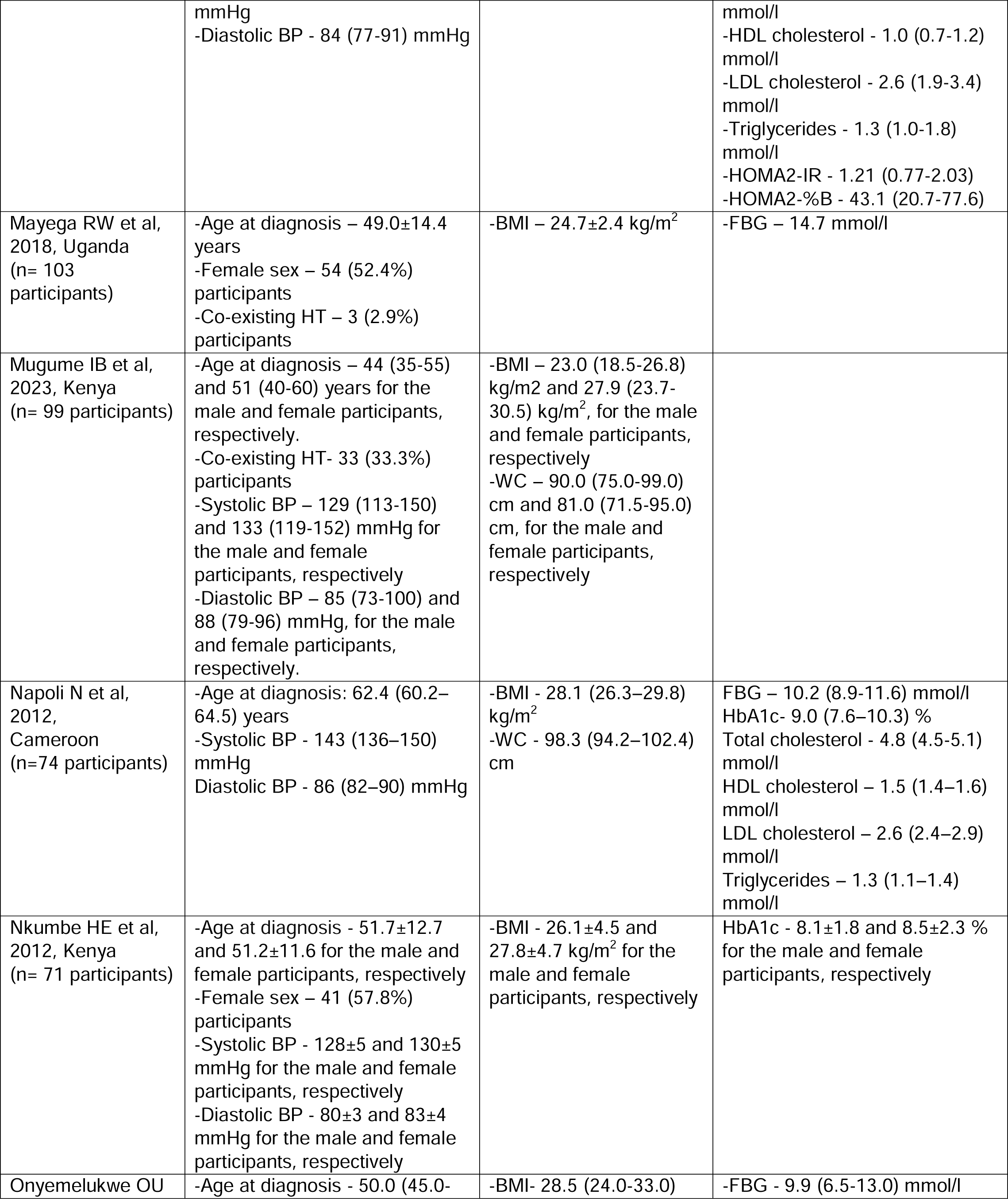
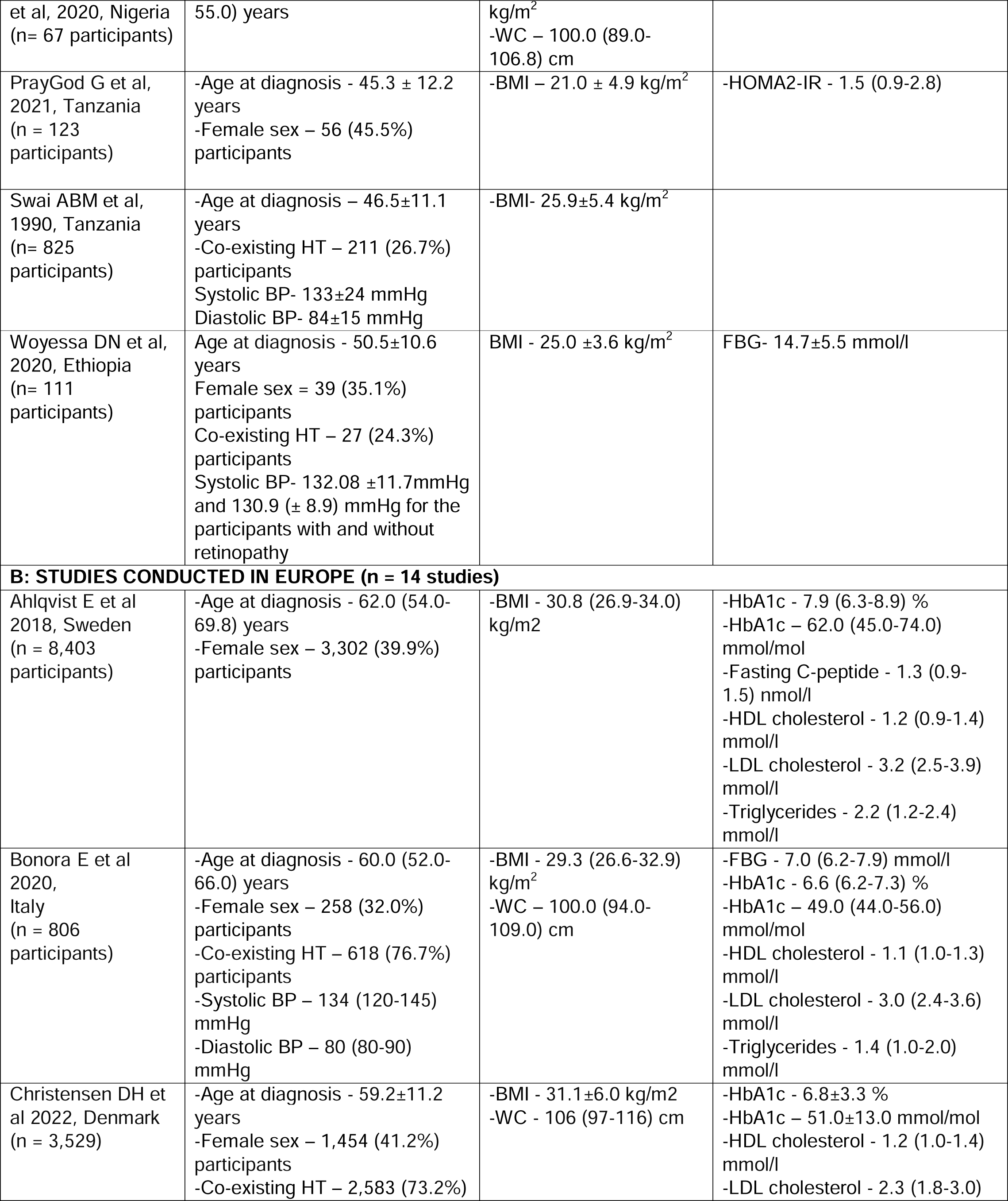
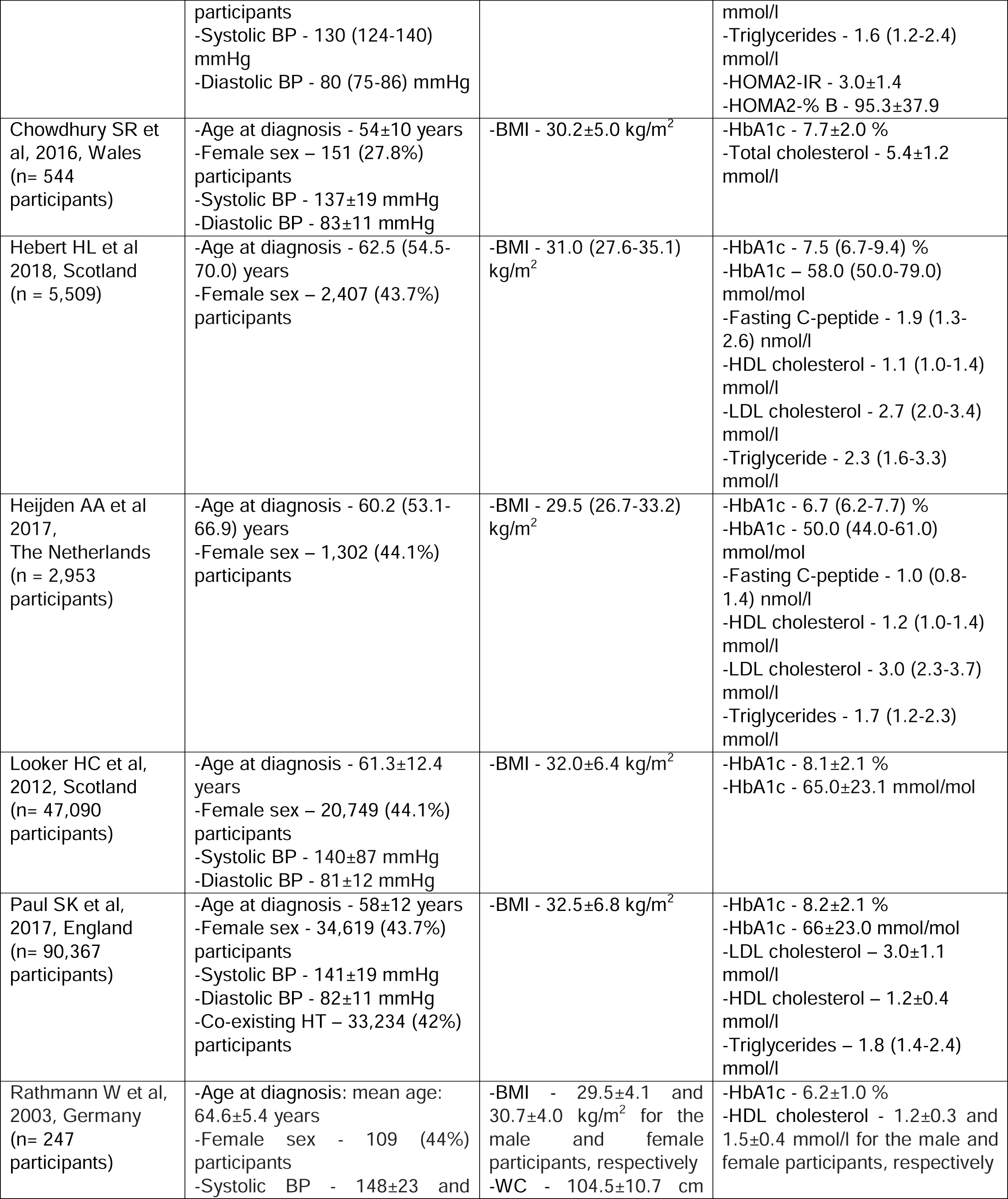
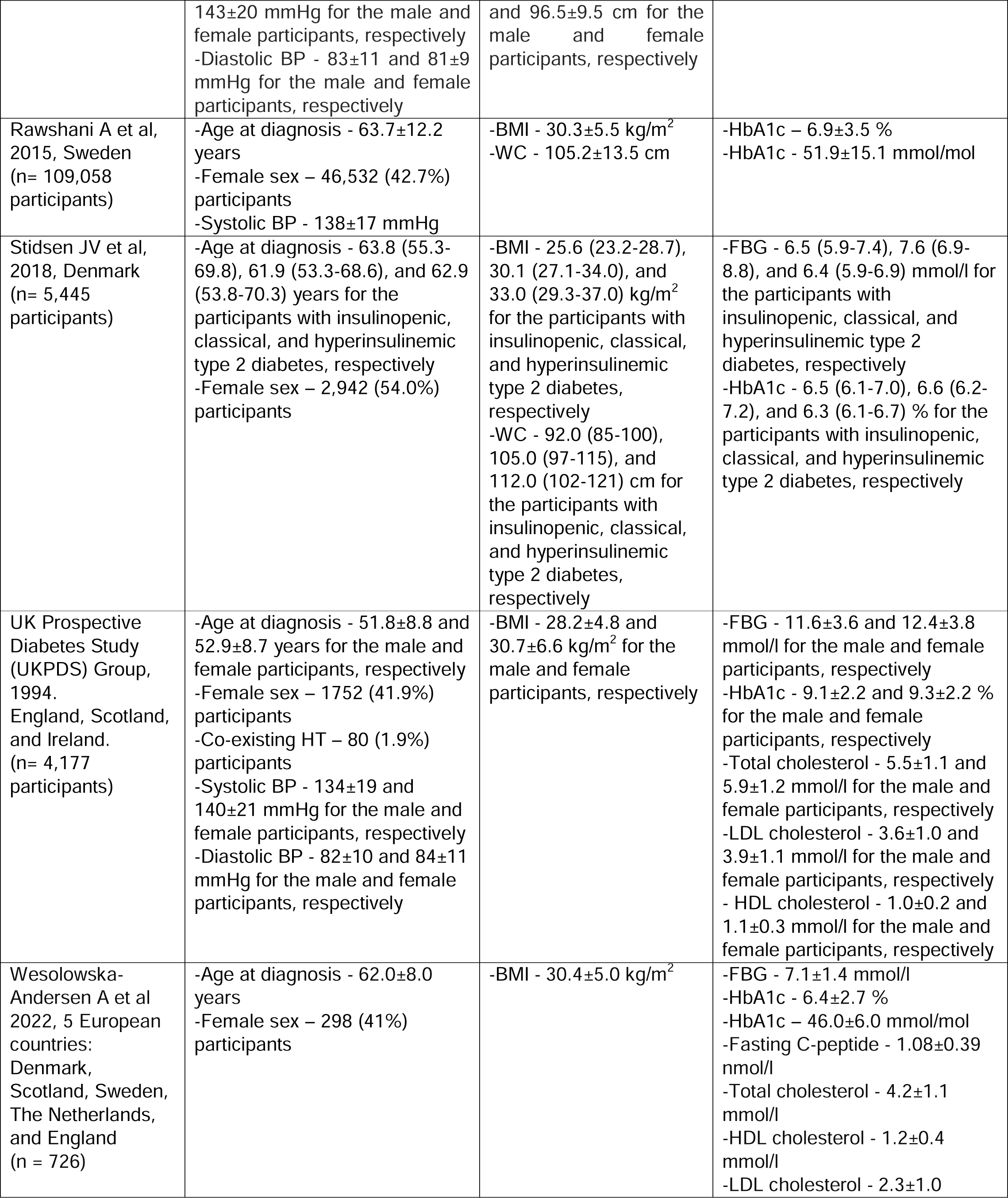
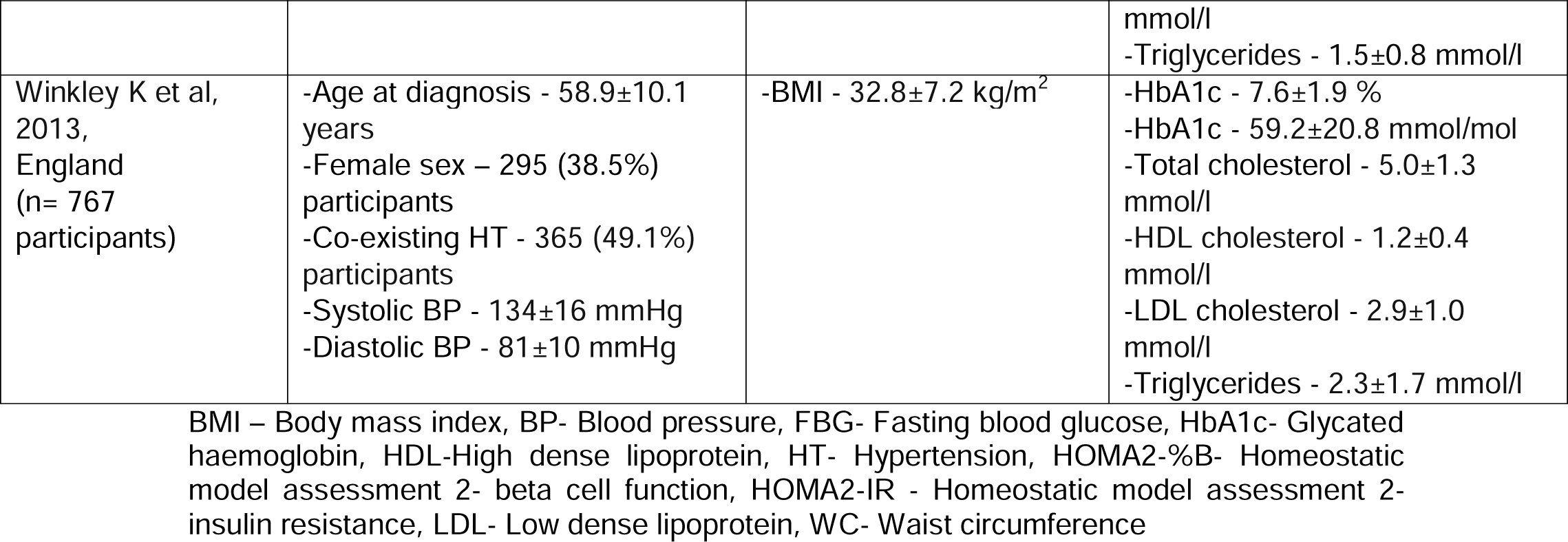
Characteristics of each study included in the systematic review.

| **1<sup>st</sup> author, publication year, country, and number of participants** | **Demographic and clinical characteristics** | **Anthropometric characteristics** | **Metabolic characteristics** |
| --- | --- | --- | --- |
| **A: STUDIES CONDUCTED IN AFRICA (n= 12)** |  |  |  |
| Amoah AG et al, 2002, Ghana (n= 300 participants) | Age at diagnosis – 52.9±14.2 years<br>Female sex – 155 (51.7) participants<br>Systolic BP- 146±30 mmHg<br>Diastolic BP- 82±14 mmHg | BMI – 26.6±6.3 kg/m<sup>2</sup> | FBG – 8.0±6.3 mmol/l |
| Ekun OA et al, 2022, Nigeria (n= 160 participants) | -Age at diagnosis – 59.0±9.0 years<br>-Female sex – 102 (63.8%) participants<br>-Systolic BP - 141±11 mmHg<br>-Diastolic BP- 89±9 mmHg | -BMI - 31.8±4.3 kg/m<sup>2</sup> | -FBG – 9.1±2.8 mmol/l<br>-HbA1c - 8.8±2.8 %<br>-Total cholesterol – 4.9±0.9 mmol/l<br>-Triglycerides – 1.2±0.4 mmol/l<br>- HDL cholesterol – 1.1±0.2 mmol/l<br>-LDL cholesterol – 3.3±0.7 mmol/l |
| Hird TR et al, 2016, South Africa (n= 153 participants) | -Age at diagnosis - 55.4 (53.4 - 57.5) years<br>-Female sex – 129 (84.3%) participants<br>-Co-existing HT- 114 (74.3%) participants<br>-Systolic BP- 128 (125–132) mmHg<br>-Diastolic BP- 83 (81–85) mmHg | -BMI - 34.6 (32.9–36.3) kg/m<sup>2</sup><br>-WC - 110.1 (107.1–113.2) cm | -FBG - 8.3 (7.6–9.0) mmol/l<br>-HbA1c - 8.0 (7.6–8.4) %<br>-Total cholesterol - 4.9 (4.8–5.1) mmol/l<br>-Triglycerides - 1.8 (1.6–2.0) mmol/l<br>-HDL cholesterol - 1.3 (1.2–1.3) mmol/l<br>-LDL cholesterol - 2.8 (2.6–2.9) mmol/l |
| Kibirige et al, 2022, Uganda, (n= 500 participants) | -Age at diagnosis - 48 (39-58) years<br>-Female sex- 283 (56.6%) participants<br>-Co-existing HT – 175 (35%) participants<br>-Systolic BP - 126 (115-137) | -BMI - 27.5 (23.6-31.4) kg/m<sup>2</sup><br>-WC - 96.0 (87.0-104.8) cm | -FBG - 8.6 (6.2-13.4) mmol/l<br>-HbA1c - 10.3 (7.7-12.5) %<br>-HbA1c – 90.0 (61.0-113.0) mmol/mol<br>-Fasting C-peptide - 0.46 (0.27-0.70) nmol/l<br>-Total cholesterol - 4.0 (3.3-5.0) |
|  | mmHg<br>-Diastolic BP - 84 (77-91) mmHg |  | mmol/l<br>-HDL cholesterol - 1.0 (0.7-1.2) mmol/l<br>-LDL cholesterol - 2.6 (1.9-3.4) mmol/l<br>-Triglycerides - 1.3 (1.0-1.8) mmol/l<br>-HOMA2-IR - 1.21 (0.77-2.03)<br>-HOMA2-%B - 43.1 (20.7-77.6) |
| Mayega RW et al, 2018, Uganda (n= 103 participants) | -Age at diagnosis – 49.0±14.4 years<br>-Female sex – 54 (52.4%) participants<br>-Co-existing HT – 3 (2.9%) participants | -BMI – 24.7±2.4 kg/m<sup>2</sup> | -FBG – 14.7 mmol/l |
| Mugume IB et al, 2023, Kenya (n= 99 participants) | -Age at diagnosis – 44 (35-55) and 51 (40-60) years for the male and female participants, respectively.<br>-Co-existing HT- 33 (33.3%) participants<br>-Systolic BP – 129 (113-150) and 133 (119-152) mmHg for the male and female participants, respectively<br>-Diastolic BP – 85 (73-100) and 88 (79-96) mmHg, for the male and female participants, respectively. | -BMI – 23.0 (18.5-26.8) kg/m<sup>2</sup> and 27.9 (23.7-30.5) kg/m<sup>2</sup>, for the male and female participants, respectively<br>-WC – 90.0 (75.0-99.0) cm and 81.0 (71.5-95.0) cm, for the male and female participants, respectively |  |
| Napoli N et al, 2012, Cameroon (n=74 participants) | -Age at diagnosis: 62.4 (60.2–64.5) years<br>-Systolic BP - 143 (136–150) mmHg<br>Diastolic BP - 86 (82–90) mmHg | -BMI - 28.1 (26.3–29.8) kg/m<sup>2</sup><br>-WC - 98.3 (94.2–102.4) cm | FBG – 10.2 (8.9-11.6) mmol/l<br>HbA1c- 9.0 (7.6–10.3) %<br>Total cholesterol - 4.8 (4.5-5.1) mmol/l<br>HDL cholesterol – 1.5 (1.4–1.6) mmol/l<br>LDL cholesterol – 2.6 (2.4–2.9) mmol/l<br>Triglycerides – 1.3 (1.1–1.4) mmol/l |
| Nkumbe HE et al, 2012, Kenya (n= 71 participants) | -Age at diagnosis - 51.7±12.7 and 51.2±11.6 for the male and female participants, respectively<br>-Female sex – 41 (57.8%) participants<br>-Systolic BP - 128±5 and 130±5 mmHg for the male and female participants, respectively<br>-Diastolic BP - 80±3 and 83±4 mmHg for the male and female participants, respectively | -BMI - 26.1±4.5 and 27.8±4.7 kg/m<sup>2</sup> for the male and female participants, respectively | HbA1c - 8.1±1.8 and 8.5±2.3 % for the male and female participants, respectively |
| Onyemelukwe OU | -Age at diagnosis - 50.0 (45.0- | -BMI- 28.5 (24.0-33.0) | -FBG - 9.9 (6.5-13.0) mmol/l |
| et al, 2020, Nigeria<br>(n= 67 participants) | 55.0) years | kg/m<sup>2</sup><br>-WC – 100.0 (89.0-106.8) cm |  |
| PrayGod G et al, 2021, Tanzania<br>(n = 123 participants) | -Age at diagnosis - 45.3 ± 12.2 years<br>-Female sex – 56 (45.5%) participants | -BMI – 21.0 ± 4.9 kg/m<sup>2</sup> | -HOMA2-IR - 1.5 (0.9-2.8) |
| Swai ABM et al, 1990, Tanzania<br>(n= 825 participants) | -Age at diagnosis – 46.5±11.1 years<br>-Co-existing HT – 211 (26.7%) participants<br>Systolic BP- 133±24 mmHg<br>Diastolic BP- 84±15 mmHg | -BMI- 25.9±5.4 kg/m<sup>2</sup> |  |
| Woyessa DN et al, 2020, Ethiopia<br>(n= 111 participants) | Age at diagnosis - 50.5±10.6 years<br>Female sex = 39 (35.1%) participants<br>Co-existing HT – 27 (24.3%) participants<br>Systolic BP- 132.08 ±11.7mmHg and 130.9 (± 8.9) mmHg for the participants with and without retinopathy | BMI - 25.0 ±3.6 kg/m<sup>2</sup> | FBG- 14.7±5.5 mmol/l |
| **B: STUDIES CONDUCTED IN EUROPE (n = 14 studies)** |  |  |  |
| Ahlqvist E et al 2018, Sweden<br>(n = 8,403 participants) | -Age at diagnosis - 62.0 (54.0-69.8) years<br>-Female sex – 3,302 (39.9%) participants | -BMI - 30.8 (26.9-34.0) kg/m<sup>2</sup> | -HbA1c - 7.9 (6.3-8.9) %<br>-HbA1c – 62.0 (45.0-74.0) mmol/mol<br>-Fasting C-peptide - 1.3 (0.9-1.5) nmol/l<br>-HDL cholesterol - 1.2 (0.9-1.4) mmol/l<br>-LDL cholesterol - 3.2 (2.5-3.9) mmol/l<br>-Triglycerides - 2.2 (1.2-2.4) mmol/l |
| Bonora E et al 2020, Italy<br>(n = 806 participants) | -Age at diagnosis - 60.0 (52.0-66.0) years<br>-Female sex – 258 (32.0%) participants<br>-Co-existing HT – 618 (76.7%) participants<br>-Systolic BP – 134 (120-145) mmHg<br>-Diastolic BP – 80 (80-90) mmHg | -BMI - 29.3 (26.6-32.9) kg/m<sup>2</sup><br>-WC – 100.0 (94.0-109.0) cm | -FBG - 7.0 (6.2-7.9) mmol/l<br>-HbA1c - 6.6 (6.2-7.3) %<br>-HbA1c – 49.0 (44.0-56.0) mmol/mol<br>-HDL cholesterol - 1.1 (1.0-1.3) mmol/l<br>-LDL cholesterol - 3.0 (2.4-3.6) mmol/l<br>-Triglycerides - 1.4 (1.0-2.0) mmol/l |
| Christensen DH et al 2022, Denmark<br>(n = 3,529) | -Age at diagnosis - 59.2±11.2 years<br>-Female sex – 1,454 (41.2%) participants<br>-Co-existing HT – 2,583 (73.2%) | -BMI - 31.1±6.0 kg/m<sup>2</sup><br>-WC - 106 (97-116) cm | -HbA1c - 6.8±3.3 %<br>-HbA1c – 51.0±13.0 mmol/mol<br>-HDL cholesterol - 1.2 (1.0-1.4) mmol/l<br>-LDL cholesterol - 2.3 (1.8-3.0) |
|  | participants<br>-Systolic BP - 130 (124-140) mmHg<br>-Diastolic BP - 80 (75-86) mmHg |  | mmol/l<br>-Triglycerides - 1.6 (1.2-2.4) mmol/l<br>-HOMA2-IR - 3.0±1.4<br>-HOMA2-% B - 95.3±37.9 |
| Chowdhury SR et al, 2016, Wales (n= 544 participants) | -Age at diagnosis - 54±10 years<br>-Female sex – 151 (27.8%) participants<br>-Systolic BP - 137±19 mmHg<br>-Diastolic BP - 83±11 mmHg | -BMI - 30.2±5.0 kg/m<sup>2</sup> | -HbA1c - 7.7±2.0 %<br>-Total cholesterol - 5.4±1.2 mmol/l |
| Hebert HL et al 2018, Scotland (n = 5,509) | -Age at diagnosis - 62.5 (54.5-70.0) years<br>-Female sex – 2,407 (43.7%) participants | -BMI - 31.0 (27.6-35.1) kg/m<sup>2</sup> | -HbA1c - 7.5 (6.7-9.4) %<br>-HbA1c – 58.0 (50.0-79.0) mmol/mol<br>-Fasting C-peptide - 1.9 (1.3-2.6) nmol/l<br>-HDL cholesterol - 1.1 (1.0-1.4) mmol/l<br>-LDL cholesterol - 2.7 (2.0-3.4) mmol/l<br>-Triglyceride - 2.3 (1.6-3.3) mmol/l |
| Heijden AA et al 2017, The Netherlands (n = 2,953 participants) | -Age at diagnosis - 60.2 (53.1-66.9) years<br>-Female sex – 1,302 (44.1%) participants | -BMI - 29.5 (26.7-33.2) kg/m<sup>2</sup> | -HbA1c - 6.7 (6.2-7.7) %<br>-HbA1c - 50.0 (44.0-61.0) mmol/mol<br>-Fasting C-peptide - 1.0 (0.8-1.4) nmol/l<br>-HDL cholesterol - 1.2 (1.0-1.4) mmol/l<br>-LDL cholesterol - 3.0 (2.3-3.7) mmol/l<br>-Triglycerides - 1.7 (1.2-2.3) mmol/l |
| Looker HC et al, 2012, Scotland (n= 47,090 participants) | -Age at diagnosis - 61.3±12.4 years<br>-Female sex – 20,749 (44.1%) participants<br>-Systolic BP - 140±87 mmHg<br>-Diastolic BP - 81±12 mmHg | -BMI - 32.0±6.4 kg/m<sup>2</sup> | -HbA1c - 8.1±2.1 %<br>-HbA1c - 65.0±23.1 mmol/mol |
| Paul SK et al, 2017, England (n= 90,367 participants) | -Age at diagnosis - 58±12 years<br>-Female sex - 34,619 (43.7%) participants<br>-Systolic BP - 141±19 mmHg<br>-Diastolic BP - 82±11 mmHg<br>-Co-existing HT – 33,234 (42%) participants | -BMI - 32.5±6.8 kg/m<sup>2</sup> | -HbA1c - 8.2±2.1 %<br>-HbA1c - 66±23.0 mmol/mol<br>-LDL cholesterol – 3.0±1.1 mmol/l<br>-HDL cholesterol – 1.2±0.4 mmol/l<br>-Triglycerides – 1.8 (1.4-2.4) mmol/l |
| Rathmann W et al, 2003, Germany (n= 247 participants) | -Age at diagnosis: mean age: 64.6±5.4 years<br>-Female sex - 109 (44%) participants<br>-Systolic BP - 148±23 and | -BMI - 29.5±4.1 and 30.7±4.0 kg/m<sup>2</sup> for the male and female participants, respectively<br>-WC - 104.5±10.7 cm | -HbA1c - 6.2±1.0 %<br>-HDL cholesterol - 1.2±0.3 and 1.5±0.4 mmol/l for the male and female participants, respectively |
|  | 143±20 mmHg for the male and female participants, respectively<br>-Diastolic BP - 83±11 and 81±9 mmHg for the male and female participants, respectively | and 96.5±9.5 cm for the male and female participants, respectively |  |
| Rawshani A et al, 2015, Sweden (n= 109,058 participants) | -Age at diagnosis - 63.7±12.2 years<br>-Female sex – 46,532 (42.7%) participants<br>-Systolic BP - 138±17 mmHg | -BMI - 30.3±5.5 kg/m<sup>2</sup><br>-WC - 105.2±13.5 cm | -HbA1c – 6.9±3.5 %<br>-HbA1c - 51.9±15.1 mmol/mol |
| Stidsen JV et al, 2018, Denmark (n= 5,445 participants) | -Age at diagnosis - 63.8 (55.3-69.8), 61.9 (53.3-68.6), and 62.9 (53.8-70.3) years for the participants with insulinopenic, classical, and hyperinsulinemic type 2 diabetes, respectively<br>-Female sex – 2,942 (54.0%) participants | -BMI - 25.6 (23.2-28.7), 30.1 (27.1-34.0), and 33.0 (29.3-37.0) kg/m<sup>2</sup> for the participants with insulinopenic, classical, and hyperinsulinemic type 2 diabetes, respectively<br>-WC - 92.0 (85-100), 105.0 (97-115), and 112.0 (102-121) cm for the participants with insulinopenic, classical, and hyperinsulinemic type 2 diabetes, respectively | -FBG - 6.5 (5.9-7.4), 7.6 (6.9-8.8), and 6.4 (5.9-6.9) mmol/l for the participants with insulinopenic, classical, and hyperinsulinemic type 2 diabetes, respectively<br>-HbA1c - 6.5 (6.1-7.0), 6.6 (6.2-7.2), and 6.3 (6.1-6.7) % for the participants with insulinopenic, classical, and hyperinsulinemic type 2 diabetes, respectively |
| UK Prospective Diabetes Study (UKPDS) Group, 1994. England, Scotland, and Ireland. (n= 4,177 participants) | -Age at diagnosis - 51.8±8.8 and 52.9±8.7 years for the male and female participants, respectively<br>-Female sex – 1752 (41.9%) participants<br>-Co-existing HT – 80 (1.9%) participants<br>-Systolic BP - 134±19 and 140±21 mmHg for the male and female participants, respectively<br>-Diastolic BP - 82±10 and 84±11 mmHg for the male and female participants, respectively | -BMI - 28.2±4.8 and 30.7±6.6 kg/m<sup>2</sup> for the male and female participants, respectively | -FBG - 11.6±3.6 and 12.4±3.8 mmol/l for the male and female participants, respectively<br>-HbA1c - 9.1±2.2 and 9.3±2.2 % for the male and female participants, respectively<br>-Total cholesterol - 5.5±1.1 and 5.9±1.2 mmol/l for the male and female participants, respectively<br>-LDL cholesterol - 3.6±1.0 and 3.9±1.1 mmol/l for the male and female participants, respectively<br>- HDL cholesterol - 1.0±0.2 and 1.1±0.3 mmol/l for the male and female participants, respectively |
| Wesolowska-Andersen A et al 2022, 5 European countries: Denmark, Scotland, Sweden, The Netherlands, and England (n = 726) | -Age at diagnosis - 62.0±8.0 years<br>-Female sex – 298 (41%) participants | -BMI - 30.4±5.0 kg/m<sup>2</sup> | -FBG - 7.1±1.4 mmol/l<br>-HbA1c - 6.4±2.7 %<br>-HbA1c – 46.0±6.0 mmol/mol<br>-Fasting C-peptide - 1.08±0.39 nmol/l<br>-Total cholesterol - 4.2±1.1 mmol/l<br>-HDL cholesterol - 1.2±0.4 mmol/l<br>-LDL cholesterol - 2.3±1.0 |
|  |  |  | mmol/l<br>-Triglycerides - 1.5±0.8 mmol/l |
| Winkley K et al, 2013, England (n= 767 participants) | -Age at diagnosis - 58.9±10.1 years<br>-Female sex – 295 (38.5%) participants<br>-Co-existing HT - 365 (49.1%) participants<br>-Systolic BP - 134±16 mmHg<br>-Diastolic BP - 81±10 mmHg | -BMI - 32.8±7.2 kg/m<sup>2</sup> | -HbA1c - 7.6±1.9 %<br>-HbA1c - 59.2±20.8 mmol/mol<br>-Total cholesterol - 5.0±1.3 mmol/l<br>-HDL cholesterol - 1.2±0.4 mmol/l<br>-LDL cholesterol - 2.9±1.0 mmol/l<br>-Triglycerides - 2.3±1.7 mmol/l |

BMI – Body mass index, BP- Blood pressure, FBG- Fasting blood glucose, HbA1c- Glycated haemoglobin, HDL-High dense lipoprotein, HT- Hypertension, HOMA2-%B- Homeostatic model assessment 2- beta cell function, HOMA2-IR - Homeostatic model assessment 2- insulin resistance, LDL- Low dense lipoprotein, WC- Waist circumference

Of the 26 studies included in this systematic review, 12 studies (46.2%) [11-22] and 14 studies (53.8%) [23-36] were conducted on 2,586 Black Africans in eight countries and 279,621 White Europeans in nine countries, respectively.

The African studies were conducted in Ghana [11], Uganda [12, 19], Kenya [13, 21], Cameroon [14], Nigeria [15, 17], Tanzania [16, 20], South Africa [18], and Ethiopia [22]. In contrast, the European studies were conducted in Sweden [23, 32], Italy [24], Denmark [25, 27], Scotland [26, 33, 36], The Netherlands [29], England [30, 33, 35], Germany [31], Ireland [33], and Wales [34]. Two studies recruited participants from more than one European country [28, 33].

Among the studies included in this systematic review, only four were case-control in design [14, 15, 17, 30] with the rest being cross-sectional [11-13, 16, 18-26, 28, 29, 31-36]. All included studies reported information on the age at diagnosis, number, and proportion of female participants, and BMI of the recruited participants. The proportion of participants with co-existing hypertension, and/or systolic, and diastolic blood pressure levels was reported in 20 studies (76.9%) [11-14, 16-22, 24, 25, 30-36]. Regarding the metabolic characteristics of the participants, 23 studies (88.5%) reported information on the HbA1c and/or FBG levels as markers of glycaemic status [11, 12, 14, 15, 17-19, 21-36]. Information on the lipid profile, fasting C-peptide, HOMA2-IR, and HOMA2-%B was provided by 15 studies (57.7%) [14, 17-19, 23-26, 28-31, 33-35], five studies (19.2%) [19, 23, 26, 28, 29], three studies (11.5%) [19, 20, 25], and two studies (7.7%) [19, 25], respectively.

### Assessment of study quality

The assessment of the quality of studies is summarised in Supplementary Tables 2 and 3.

Eight (30.8%) of the 26 studies included in the systematic review were of good quality (five studies from SSA and three from Europe) [13-15, 17, 20, 23, 29, 35]. The remaining 18 studies were considered of satisfactory quality (seven studies from SSA and 11 from Europe) [11, 12, 16, 18, 19, 21, 22, 24-28, 30-34, 36].

### Assessment of the publication bias of the studies

There was no significant publication bias in the meta-analysis for the pooled prevalence of female patients in black Africans (Egger test: p=0.334) and white Europeans (Egger test: p=0.251). Similarly, studies reporting co-existing hypertension exhibited no publication bias, for both black Africans (Egger test: p=0.387) and white Europeans (Egger test: p=0.215). The funnel plots for both variables showed no substantial asymmetry (**Supplementary Figures 1 and 2**).

### Demographic and clinical characteristics of the Black Africans and White Europeans with recently diagnosed type 2 diabetes

Tables 1, 2, Figures 2, and 3 summarise the overall and specific demographic, clinical, anthropometric, and metabolic characteristics of both populations.

**Figure 2.**
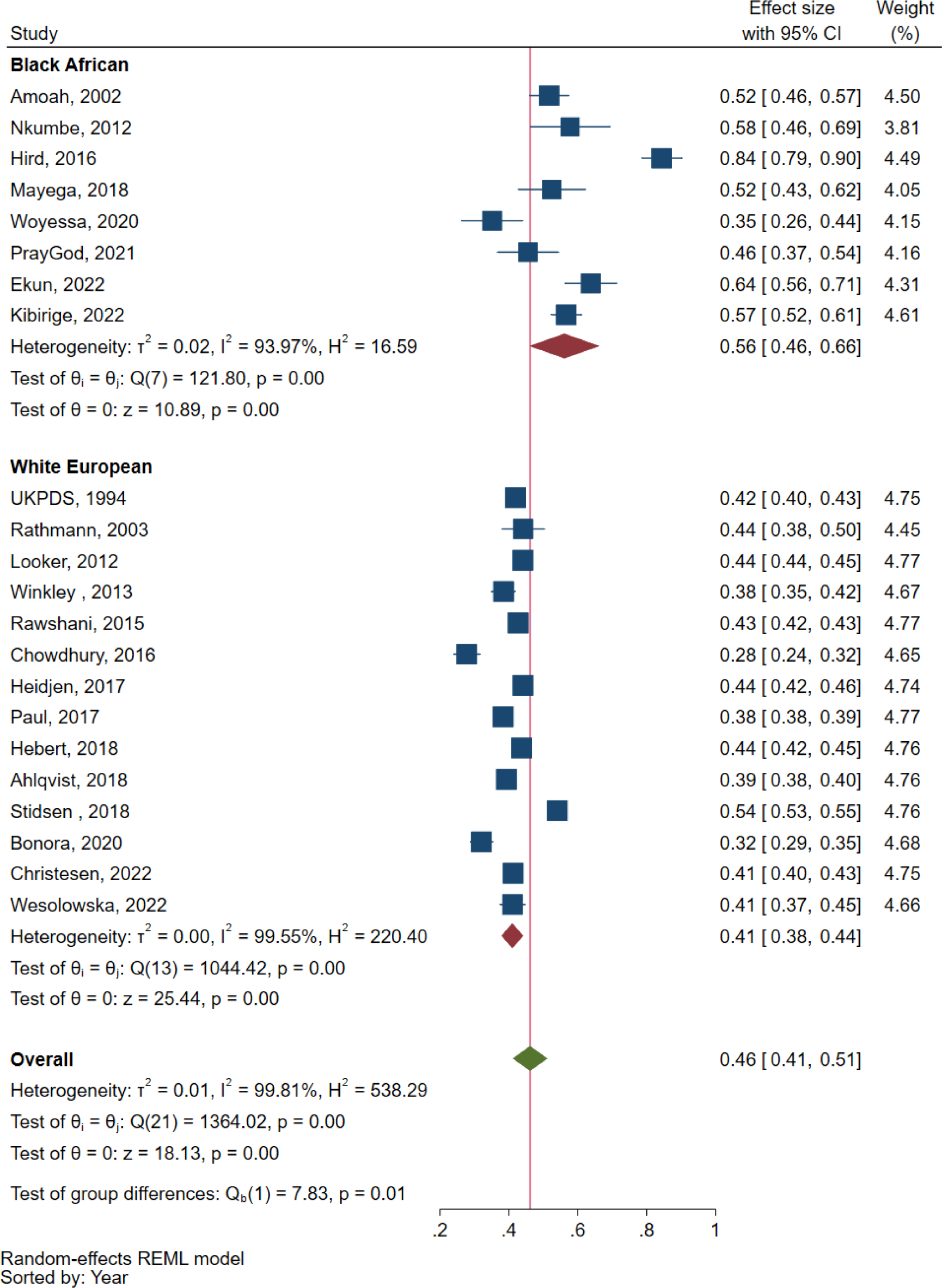
Forest plot showing the proportion of female participants in the included studies.

Compared with White Europeans, Black Africans had a lower pooled mean age at diagnosis of T2D (49.4±4.4 years vs. 61.3±2.7 years). In the Black Africans, the mean age ranged from 45.3±12.2 years in one study conducted in Tanzania [20] to 59.0±9.0 years in one study conducted in Nigeria [17]. In contrast, in the White Europeans, the mean age ranged from 51.8±8.8 years in male participants in the UK Prospective Diabetes Study (UKPDS) [33] to 64.6±5.4 years in one study conducted in Germany [31].

Studies conducted in Africa reported a higher pooled proportion of female participants than those conducted in Europe (56.0%, 95% CI 46.0%-66.0%, I^2^= 93.97%, p<0.001 vs. 41.0%, 95% CI 38%-44.1%, I^2^= 99.55%, p<0.001). Black Africans also had lower pooled proportions of self-reported history of hypertension at diagnosis of T2D compared with White Europeans (33.0%, 95% CI 14.0%-51.0%, I^2^= 98.97%, p<0.001 vs. 47.0%, 95% CI 21.0%-74.0%, I^2^= 99.96%, p<0.001) (Figures 2 **and 3**).

**Figure 3.**
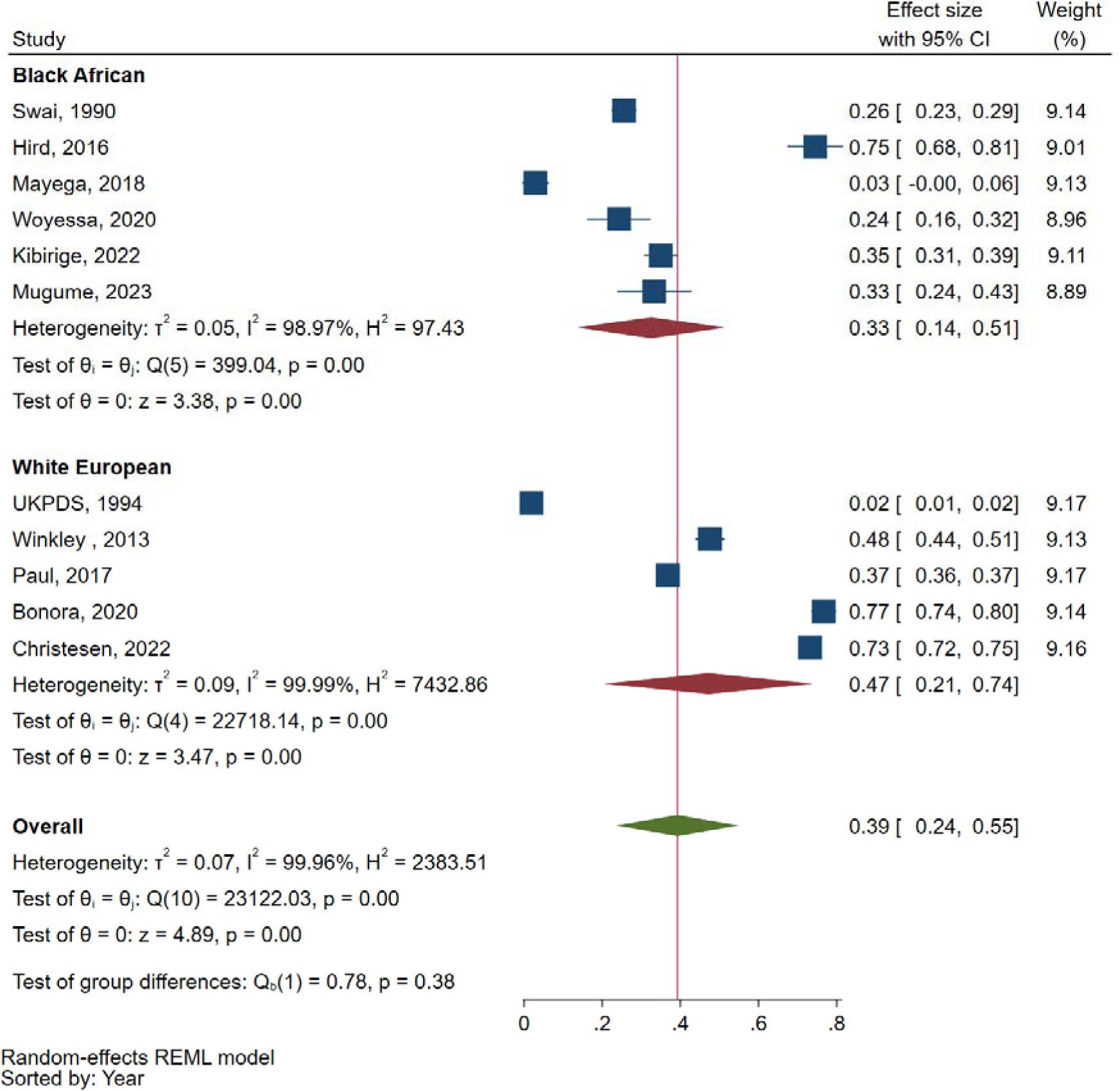
Forest plot showing the prevalence of self-reported history of hypertension in the included studies.

### Anthropometric and metabolic characteristics of the Black Africans and White Europeans with recently diagnosed type 2 diabetes

Concerning the anthropometric characteristics, compared with White Europeans, Black Africans had a lower pooled mean BMI (26.1±2.6 kg/m^2^ vs. 31.4±1.1 kg/m^2^) and median WC (99.2 [97.2-105.1] cm vs. 103.0 [100.0-106.0] cm). The mean BMI levels in Black Africans ranged from 21.0±4.9 kg/m^2^ in one study conducted in Tanzania [20] to 31.8 kg/m^2^ in one study conducted in Nigeria [17]. In the White Europeans, the mean BMI levels ranged from 28.2±4.8 kg/m^2^ in the male participants in the UKPDS [33] to 32.8±7.2 kg/m^2^ in one study conducted in England [35]. Only the Nigerian [17] and South African [18] participants had BMI levels comparable with the White European participants.

The median WC levels in Black Africans ranged from 81.0 (71.5-95.0) cm in female participants in a study by Mugume et al conducted in Kenya [13] to 110.1 (107.1-113.2) cm in a study by Hird et al conducted in South Africa [18]. In the White Europeans, the median WC participants ranged from 92.0 (85.0-100.0) cm to 112.0 (102.0-121.0) cm in the Danish participants with insulinopenic and hyperinsulinemic type 2 diabetes phenotypes, respectively [27].

Differences in lipid profile were also noted between both populations. Compared with White Europeans, Black Africans had lower pooled median LDLC (2.6 [2.5-2.8] mmol/l vs. 3.0 [2.9-3.1] mmol/l) and triglyceride (1.3 [1.2-1.8] mmol/l vs. 1.7 [1.4-1.8] mmol/l) concentrations.

Black Africans presented with higher pooled median HbA1c concentrations compared with White Europeans (9.0% [8.0-10.3]% vs. 7.1 [6.7-7.7]% or 75.0 [64.0-89.0] mmol/mol vs. 54.0 [50.0-61.0] mmol/mol). The median HbA1c concentrations in Black Africans ranged from 8.0 (7.6-8.4)% or 64.0 (60.0-64.0) mmol/mol in a study conducted in South Africa [18] to 10.3 (7.7-12.5)% or 90.0 (61.0-113.0) mmol/mol in one study conducted in Uganda [19]. In contrast, the median HbA1c in White Europeans ranged from 6.3 (6.1-6.7)% or 45.0 (43.0-50.0) mmol/mol in hyperinsulinaemic Danish participants [27] to 7.9 (6.3-8.9)% or 63.0 (45.0-74.0) mmol/mol in one study conducted in Sweden [23].

Regarding the markers of pancreatic beta-cell function status reported by five studies, Ugandan participants in a study by Kibirige et al had lower median fasting C-peptide concentration (0.46 [0.27-0.70] nmol/l) [19] compared with the Dutch [29], Swedish [23], and Scottish [26] participants (pooled median C-peptide level-1.3 [1.0-1.9] nmol/l). Additionally, Ugandan participants had a lower HOMA2-%B level [19] when compared with Danish participants in a study by Christensen et al [25]. Concerning the insulin resistance status, Ugandan [19] and Tanzanian [20] participants had lower levels of HOMA2-IR when compared with Danish participants in one study [25] (1.21 [0.77-2.03] and 1.50 [0.90-2.80], vs 3.00 ± 1.40, respectively).

## DISCUSSION

This is, to our knowledge, the first systematic review to investigate the differences in the demographic, clinical, anthropometric, and metabolic characteristics between Black Africans and White Europeans with recently diagnosed T2D. In this systematic review, we report that compared with White Europeans, Black Africans at diagnosis of T2D were younger with a female predominance, and presented with less self-reported history of hypertension, lower surrogate markers of adiposity, and higher levels of hyperglycaemia. Pathophysiologically, Ugandan and Tanzanian participants presented with a greater degree of pancreatic beta-cell secretory dysfunction and less insulin resistance when compared with four White European populations.

Remarkably, lower mean age at the time of diagnosis of T2D has also been reported in individuals of African ancestry with T2D living in the UK. Analysis of the UK primary healthcare data showed that Black Africans develop T2D at a mean age of 48±12 years compared with 58±12 years in White Europeans, reflecting the onset of T2D about 10 years earlier [30]. The reasons for a lower mean age at the time of diagnosis of T2D in adult Africans are unclear. Environmental exposures and genetic factors may play a role. For example, sub-Saharan Africa (SSA) experiences a high burden of infectious diseases such as tuberculosis and HIV which, through direct effects like inducing a pro-inflammatory state or as a result of treatment of the infection (e.g. metabolic side-effects of antiretroviral therapy) may increase diabetes risk [6, 37]. Similarly, nutritional exposure may be important. Early-life (in-utero and/or early childhood) chronic malnutrition is highly prevalent in most parts of SSA, and this has been associated with the long-term risk of early-onset diabetes which has led to the concept of developmental origins of health and disease (DOHaD) [38, 39].

The higher proportion of female participants and lower prevalence of self-reported co-existing hypertension in Black Africans compared with White Europeans may broadly reflect gender-based differences in health-seeking behaviour and low screening rates for hypertension commonly observed in SSA [40-42]. Most studies conducted in SSA have reported that African women have better health-seeking behaviour compared with men, who habitually seek medical care late when most conditions are in advanced stages [40, 42]. A high proportion of undiagnosed hypertension has been widely reported in clinic-and population-based studies conducted in SSA. In one systematic review and meta-analysis of 33 studies investigating the burden of undiagnosed hypertension in 110,414 adult Africans with hypertension, a small proportion of participants were aware of their hypertensive status (pooled prevalence of 27% [95% CI 23-31]) [41].

Most Africans tend to develop T2D at lower levels of BMI compared with White Europeans in high-income countries. This was confirmed by studies conducted in Ghana [11], Uganda [12, 19], Cameroon [14], Tanzania [16, 20], Kenya [13, 21], and Ethiopia [22]. Furthermore, people of African ancestry living in the UK have also been reported to develop T2D at a lower BMI compared with White Europeans [43]. However, participants in one study conducted in Nigeria [17] and South Africa [18] had comparable BMI levels with White European participants. This is likely to reflect differences in the stages of socioeconomic and nutrition transitions among some African countries, although other factors cannot be ruled out.

Nonetheless, the finding of a lower BMI in some African populations compared with White Europeans has potential implications for T2D prevention and management strategies. For example, we do not know whether lifestyle interventions to reduce body weight will work as well in African individuals with T2D and low BMI. In addition, low BMI in a patient with T2D may be a predictor of pancreatic beta-cell dysfunction, an insulin-deficient phenotype, and the need for early insulin initiation [8, 44]. Indeed, Ugandan participants had metabolic features consistent with a predominance of pancreatic beta-cell secretory dysfunction rather than insulin resistance (low fasting C-peptide, HOMA2-%B, and HOMA2-IR) [19]. The observation of low pancreatic beta-cell function in Africans is also in accord with studies conducted in Black Africans living in the USA and UK [45, 46]. Furthermore, this observed low pancreatic beta-cell function may well explain the high levels of hyperglycaemia that are noted at diagnosis of T2D in African patients, although late presentation or delayed detection of T2D may also be a factor.

Given the well-elaborated differences in the manifestation of T2D in Black Africans and White Europeans, it may not be prudent to directly extrapolate the European clinical guidelines for preventing and managing T2D to some Black African populations. The selection of optimal therapies for managing T2D should be based on an in-depth understanding of the prevalent phenotype and genotype [47, 48]. Because of a clinical profile of lower surrogate markers of adiposity and a predominance of pancreatic beta-cell dysfunction in some Black African populations at diagnosis of T2D, insulin, dipeptidyl peptidase IV (DPP IV) inhibitors, or sulfonylureas would be the appropriate first-line diabetes therapies in individuals with pancreatic beta-cell secretory function, with metformin best suited as first line for those with an insulin-resistant phenotype [48]. We, however, currently lack solid evidence that necessitates rigorous clinical trials to investigate the optimal prevention and management of the T2D phenotypes in Black Africans.

This systematic review had its strengths and limitations. To our knowledge, it is the first systematic review to explore the phenotypic differences between Black Africans and White Europeans with recently diagnosed T2D. Among its limitations is the small number of participants with recently diagnosed diabetes and limited information on the metabolic characteristics (notably markers of pancreatic beta-cell function and insulin resistance) in the African studies.

## CONCLUSION

This systematic review demonstrates that T2D manifests differently in Black Africans and White Europeans. This underscores the need to understand in great detail the underlying genetic and environmental factors influencing these phenotypic differences and also undertake rigorous interventional studies to provide compelling local evidence of how to optimally manage and prevent T2D in Black African populations.

## Data Availability

All data produced in the present work are contained in the manuscript.

## Acknowledgements

None.

## Availability of data

No data set is available for this systematic review. The studies included in this systematic review are published and freely available online.

## Contributorship statement

DK, WL, and MJN conceived the idea of performing this systematic review, DK, APK, and FB screened the retrieved published articles and identified the studies to include in the systematic review, BM performed the database search, APK assessed the quality of the included studies, RO performed the statistical analysis, DK extracted the key information of interest from the identified studies and wrote the initial draft of this manuscript, WL and FB provided additional interpretation of the extracted data, and MJN supervised this work. All the authors reviewed the different versions of the manuscript and read and approved the final draft of the manuscript.

