## Supplementary Tables and Figures for "Differential manifestation of type 2 diabetes in Black Africans and White Europeans with recently diagnosed type 2 diabetes: A systematic review": Supplementary Figures 1 and 2-Funnel plots.docx

**Supplementary Figure 1. Funnel plot for the studies reporting the proportion of female participants in the Black African and White European populations**

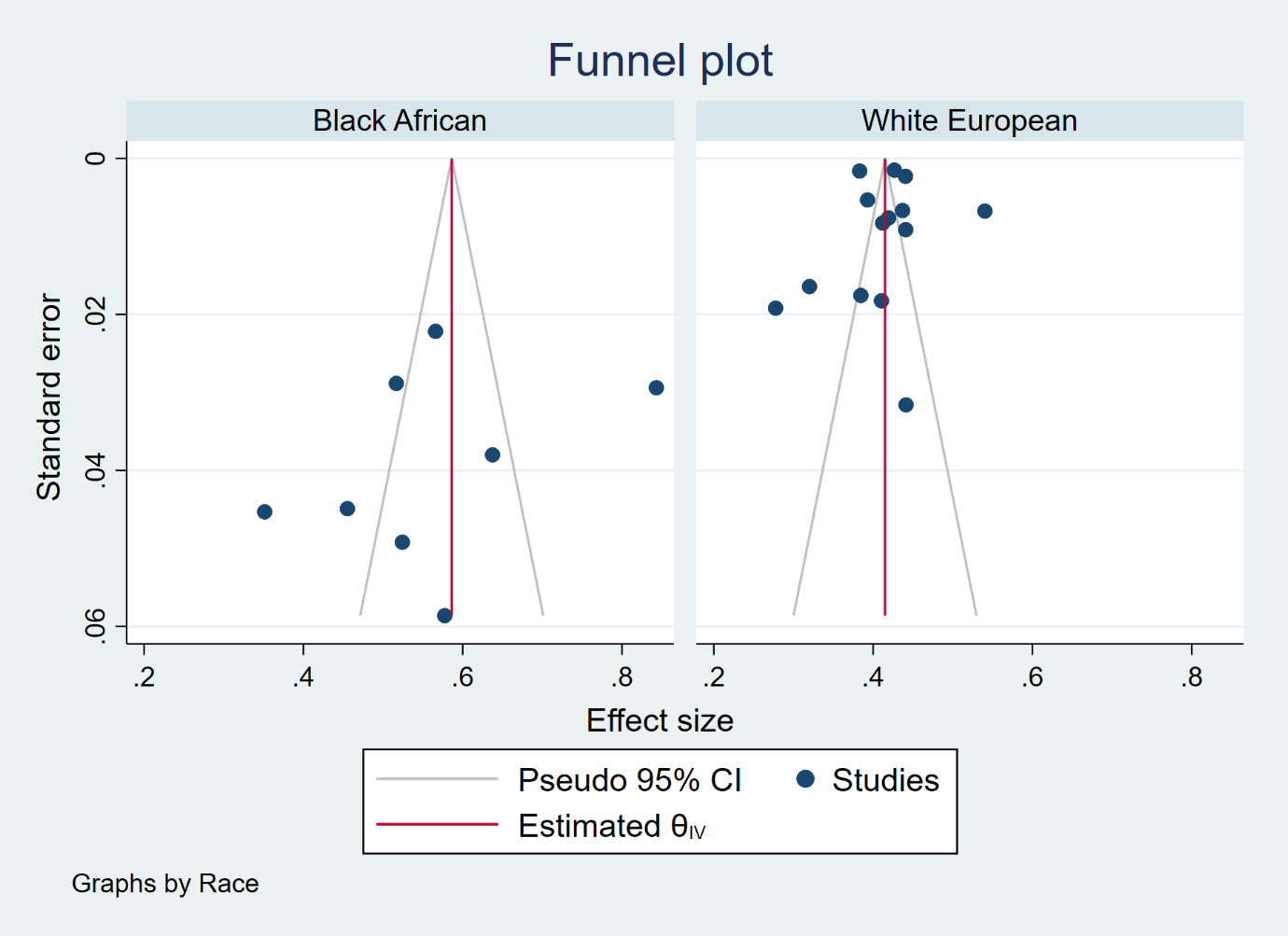

**Supplementary Figure 2. Funnel plot for the studies reporting the prevalence of self-reported history of hypertension in the Black African and White European populations**

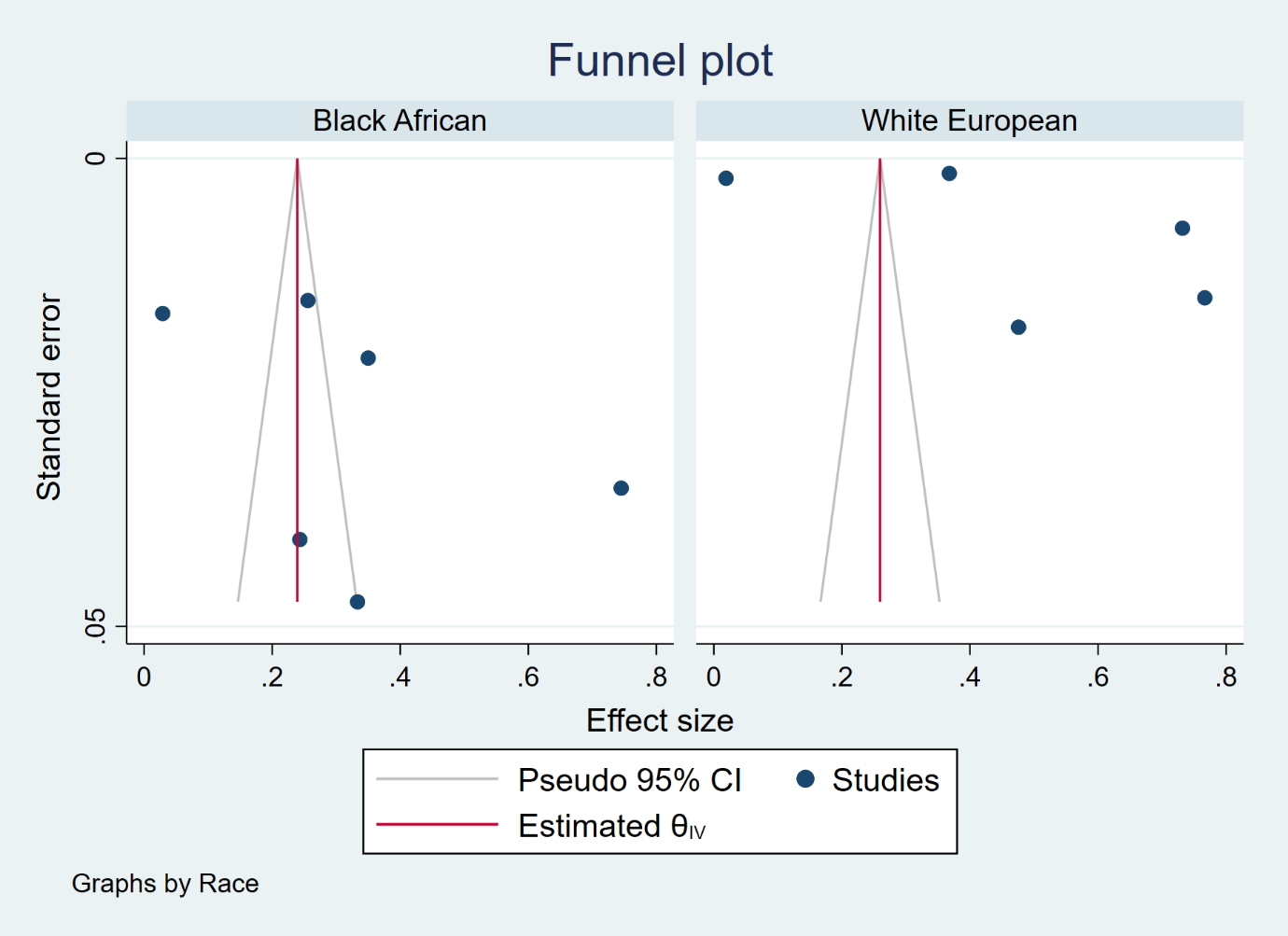
