## Supplementary Tables and Figures for "Differential manifestation of type 2 diabetes in Black Africans and White Europeans with recently diagnosed type 2 diabetes: A systematic review": Supplementary Table 3. Study quality assessment-DM SYST RVV- Europe.docx

**Supplementary Table 2. Quality of the studies included in the systematic review- Europe**

| **AUTHOR, YEAR** | | Christensen et al, 2022 | Paul et al, 2017 | Ahlqvist et al, 2018 | Hebert et al, 2017 | Heijden et al, 2017 | Wesolowska-Andersen A et al. 2022 | Chowdhury et al, 2016 | Stidsen et al | Looker et al, 2012 | Bonora et al, 2020 | Winkley et al, 2013 | UK prospective diabetes study group | Rathmann et al, 2003 | Rawshani et al, 2015 |
| --- | --- | --- | --- | --- | --- | --- | --- | --- | --- | --- | --- | --- | --- | --- | --- |
| **CRITERIA** | |  |  |  |  |  |  |  |  |  |  |  |  |  |  |
|  | **SELECTION** (Max 5 stars) | | | | | | | | | | | |  |  |  |
| **1)** **Representativeness of the sample**  a) Truly representative of the average in the target population. * (all subjects or random sampling)  b) Somewhat representative of the average in the target population. * (non-random sampling)  c) Selected group of users.  d) No description of the sampling strategy. | | 1 | 1 | 1 | 1 | 1 | 1 | 1 | 1 | 1 | 1 | 1 | 1 | 1 | 1 |
| **2) Sample size**  a) Justified and satisfactory. *  b) Not justified | | 0 | 0 | 1 | 0 | 1 | 0 | 0 | 0 | 0 | 0 | 0 | 0 | 0 | 0 |
| **3) Non-respondents**  a) Comparability between respondents and non-respondents’ characteristics is established, and the response rate is satisfactory. *  b) The response rate is unsatisfactory, or the comparability between respondents and non-respondents is unsatisfactory.  c) No description of the response rate or the characteristics of the responders and the non-responders. | | 0 | 0 | 0 | 0 | 1 | 0 | 0 | 0 | 0 | 0 | 1 | 0 | 0 | 0 |
| **4) Ascertainment of the exposure (risk factor)**  a) Validated measurement tool. **  b) Non-validated measurement tool, but the tool is available or described. *  c) No description of the measurement tool. | | 2 | 1 | 2 | 2 | 2 | 2 | 2 | 1 | 1 | 2 | 2 | 1 | 2 | 1 |
|  | **COMPARABILITY** (Max 2 stars) | | | | | | | | | | | |  |  |  |
| **1) The subjects in different outcome groups are comparable, based on the study design or analysis. Confounding factors are controlled.**  a) The study controls for the most important factor (select one). *  b) The study control for any additional factor. * | | n/a | 1 | n/a | n/a | n/a | n/a | 0 | n/a | n/a | n/a | n/a | n/a | n/a | n/a |
|  | **OUTCOME** (Max 3 stars) | | | | | | | | | | | |  |  |  |
| **1) Assessment of the outcome**  a) Independent blind assessment. **  b) Record linkage. **  c) Self report. *  d) No description. | | 2 | 2 | 2 | 2 | 2 | 2 | 2 | 2 | 2 | 2 | 2 | 2 | 2 | 2 |
| **2) Statistical test**  a) The statistical test used to analyze the data is clearly described and appropriate, and the measurement of the association is presented, including confidence intervals and the probability level (p value). *  b) The statistical test is not appropriate, not described or incomplete. | | 1 | 1 | 1 | 1 | 0 | 1 | 1 | 1 | 1 | 1 | 1 | 1 | 1 | 1 |
| **TOTAL** | | 6 | 6 | 7 | 6 | 7 | 6 | 6 | 5 | 5 | 6 | 7 | 5 | 6 | 5 |

Very Good Studies: 9-10 points

Good Studies: 7-8 points

Satisfactory Studies: 5-6 points

Unsatisfactory Studies: 0 to 4 points

$- no data tool as it was a data extraction
